# SARS-CoV-2 infections in 171 countries and over time

**DOI:** 10.1101/2020.12.01.20241539

**Authors:** Stilianos Louca

## Abstract

Understanding the dynamics of the COVID-19 pandemic, evaluating the efficacy of past and current control measures, and estimating vaccination needs, requires knowledge of the number of infections in the population over time. This number, however, generally differs substantially from the number of confirmed cases due to a large fraction of asymptomatic infections as well as geographically and temporally variable testing effort and strategies. Here I use age-stratified death count statistics, age-dependent infection fatality risks and stochastic modeling to estimate the prevalence and growth of SARS-CoV-2 infections among adults (age ≥ 20 years) in 171 countries, from early 2020 until April 9, 2021. The accuracy of the approach is confirmed through comparison to previous nationwide general-population seroprevalence surveys in multiple countries. Estimates of infections over time, compared to reported cases, reveal that the fraction of infections that are detected vary widely over time and between countries, and hence comparisons of confirmed cases alone (between countries or time points) often yield a false picture of the pandemic’s dynamics. As of April 9, 2021, the nationwide cumulative SARS-CoV-2 prevalence (past and current infections relative to the population size) is estimated at 61% (95%-CI 42-78) for Peru, 58% (39–83) for Mexico, 57% (31–75) for Brazil, 55% (34–72) for South Africa, 29% (19-48) for the US, 26% (16–49) for the United Kingdom, 19% (12–34) for France, 19% (11–33) for Sweden, 9.6% (6.5–15) for Canada, 11% (7–19) for Germany and 0.67% (0.47–1.1) for Japan. The presented time-resolved estimates expand the possibilities to study the factors that influenced and still influence the pandemic’s progression in 171 countries. Regular updates are available at: www.loucalab.com/archive/COVID19prevalence

## Introduction

Accurate estimates of the prevalence of SARS-CoV-2 in a population are needed for evaluating disease control policies and testing strategies, determining seasonal effects, predicting future disease spread, assessing the risk of foreign travel, and determining vaccination needs [1]. Even if a retreat of the epidemic seems within reach in many countries, the efficacy of control measures in 2020 and 2021 and the environmental/political/societal factors that influenced the epidemic’s progression in each country will undoubtedly be the topic of scholarly work for years to come. Due to the existence of a large fraction of asymptomatic cases, as well as variation in reporting, testing effort and testing strategies (e.g., random vs symptom-triggered), confirmed case counts cannot be directly converted to infection counts and a comparison of confirmed case counts between countries is generally of limited informative value [2]. While large-scale seroprevalence surveys (e.g., using antibody tests) can yield information on the disease’s prevalence in a population, such surveys involve substantial financial and logistical challenges and only yield prevalence estimates at a specific time point.

In contrast to case reports, COVID-19-related death counts are generally regarded as less sensitive to testing effort and strategy [3, 4], and fortunately most countries have established nationwide continuous reporting mechanisms for death counts. Hence, in principle, knowing the infection fatality risk (IFR, the probability of death following infection by SARS-CoV-2) should permit a conversion of death counts to infection counts [3, 4]. The IFR of SARS-CoV-2, however, depends strongly on the patient’s age, and hence the effective IFR of the entire population depends on the population’s age structure as well as the disease’s age distribution [5]. Indeed, it was shown that the age-dependency of the IFR, the age-dependency of SARS-CoV-2 prevalence, and the age structure of the population are largely sufficient to explain variation in the effective IFR between countries [6]. This suggests that age-stratified death counts can (and must) be used with age-dependent IFR estimates in order to obtain an accurate estimate of infection counts. This approach has been successfully used to estimate SARS-CoV-2 prevalence over time in Europe until May 4, 2020 [4].

Unfortunately, the ongoing pandemic necessitates continuously updated prevalence estimates. Moreover, age-stratified and time-resolved death statistics are not readily available for many countries with insufficiently comprehensive reporting, thus preventing a direct adoption of the above approach [4, 7]. In cases where only total death counts are available (e.g., as disseminated by the World Health Organization) one needs to somehow independently determine the likely age distribution of infections in order to convert total death counts to infection counts. Here I address this challenge by leveraging information on the age distribution of SARS-CoV-2 infections from multiple countries with available age-stratified death reports, to estimate the likely age-distribution of SARS-CoV-2 in other countries, while accounting for each country’s age structure. Based on these calibrations, I estimate the prevalence of SARS-CoV-2 (cumulative number of infections, weekly new infections and exponential growth rate) over time in 171 countries up until April 9, 2021, among adults aged 20 years or more. My predictions are largely consistent with data from multiple previously published nationwide seroprevalence surveys.

### Calibrating the age distribution of SARS-CoV-2 prevalence

In order to calculate infection counts solely from total (i.e., non-age-stratified) death counts, while accounting for the age-dependency of the IFR and each country’s population age structure, independent estimates of the ratios of infection risks between age groups (i.e., the risk of infection in any one age group relative to any other age group) are needed. To determine the general distribution of age-specific infection risk ratios, I analyzed weekly age-stratified COVID-19-related death reports from 24 countries around the world using a probabilistic model of Poisson-distributed time-delayed death counts (see Methods for details). Briefly, for any given country *c*, any given week *w*, and any given age group *g*, I assumed that the number of new infections during that week (*I*_*c,w,g*_) is approximately equal to *α*_*c,g*_*I*_*c,w,r*_*N*_*c,g*_*/N*_*c,r*_, where *r* represents some fixed reference age group, *N*_*c,g*_ is the population size of age group *g*, and *α*_*c,g*_ is the relative risk of an individual in age group *g* being infected compared to that of an individual in age group *r*. The expected number of deaths in each age group 4 weeks later (roughly the average time lag between infection and death [8]), denoted *D*_*c,w*+4,*g*_, was assumed to be *I*_*c,w,g*_*R*_*g*_, where *R*_*g*_ is the IFR for that age group. Note that while *R*_*g*_ could in principle also vary between countries, to date insufficient information is available for calibrating *R*_*g*_ separately for each country (but see discussion of caveats below). Age-specific IFRs were calculated beforehand by taking the average over multiple IFR estimates reported in the literature [6, 7, 9–12]. This calibration thus accounts for the age-structure of each country, the age-distribution of the disease in each country and the age-dependency of the IFR. A critical assumption of the model is that, in any given country, nationwide age-specific infection risks co-vary linearly between age groups over time, i.e., an increase of disease prevalence in one age group coincides with a proportional increase of prevalence in any other age group. This assumption is motivated by the observation that nationwide death rates generally covary strongly linearly between age groups (Fig. 1A and Supplemental Fig. S1); the adequacy of this model is also confirmed in retrospect (see below). For each country, I fitted the infection risk ratios *α*_*c,g*_ (for all *g* ≠ *r*) as well as the weekly infections in the reference age-group *I*_*c,w,r*_ (one per week) to the age-stratified weekly death counts using a maximum-likelihood approach and assuming that weekly death counts follow a Poisson distribution. This stochastic model explained the data generally well, with observed weekly death counts almost always falling within the 95% confidence interval of the model’s predictions (Supplemental Fig. S2). This supports the initial assumption that infection risks co-vary approximately linearly between age groups over time and suggests that country-specific but time-independent infection risk ratios are largely sufficient for describing the age-distribution of SARS-CoV-2 infections in a country and over time. For any given age group *g*, the fitted infection risk ratios *α*_*c,g*_ differed between countries but were generally within the same order of magnitude (Fig. 1B). On the basis of this observation, and as explained in the next section, it thus seems possible to approximately estimate the number of infections in any other country based on total death counts, the population’s age structure and the pool of infection risk ratios *α*_*c,g*_ fitted above (accounting for the inevitable uncertainty in the latter).

**Figure 1:**
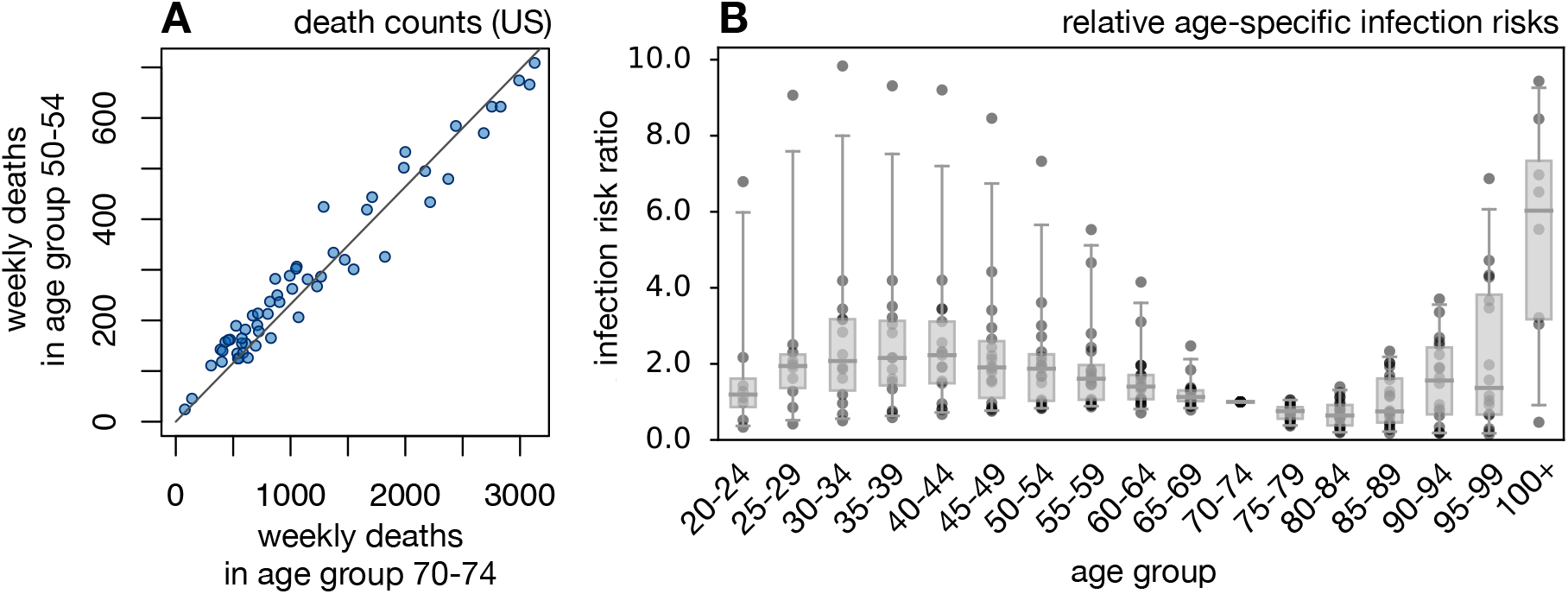
Infection and death rates covary linearly between age-groups. (A) Weekly reported COVID-19-related death counts in the US, in age group 70–74 (horizontal axis) and age group 50–54 (vertical axis). Each point corresponds to a different week (defined here as a 7-day period). The linear regression line is shown for reference. For additional age groups and countries see Supplemental Fig. S1. The strong co-linearity of death rates between age-groups suggests that infection risks also covary linearly between age-groups. (B) Relative infection risk ratios (relative to age group 70–74) for different countries, estimated based on death-stratified COVID-19-related death counts. Each column represents a different age group, and in each column each point represents a distinct country. Horizontal bars represent medians and boxes span 50%-percentiles of the data.

### Estimating infection counts over time

Based on the ensemble of fitted infection risk ratios, the same age-dependent IFRs used above, the probability distribution of time lags between infection, disease onset and death [8], and total (non-age-stratified) COVID-19-related death count reports disseminated by the WHO, I estimated the weekly infection counts over time in each of 171 countries (details in Methods). Briefly, for any given country *c*, week *w* and any given set of relative infection risks *α*_1_, *α*_2_,.., the total number of deaths during that week (*D*_*c,w*_) was assumed to be Poisson-distributed with expectation equal to:

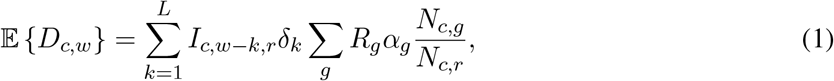

where as before *R*_*g*_ is the IFR for age group *g, N*_*c,g*_ is the population size of age group *g, δ*_*k*_ is the probability that a fatal infection will result in death after *k* weeks, *L* is the maximum considered time lag between infection and death, and *I*_*c,w,r*_ is the (a priori unknown) number of new infections in the reference age group *r* during week *w*. For the second sum in Eq. (1), I considered only age groups at 20 years or older (in 5-year intervals), because estimates of the infection risk ratios *α*_*g*_ were unreliable for younger ages (due to low death counts) and because deaths among less-than-20-year olds were numerically negligible compared to the total number of deaths reported. Note that the expected number of deaths in any given week depends on the number of infections in multiple previous weeks, due to the variability of the time lag between infection and death (typically 2–6 weeks [8]). Hence, the time series of observed weekly death counts (*D*_*c*,1_, *D*_*c*,2_,..) results from a *convolution* (“blurring”) of the weekly infections counts (*I*_*c*,1,*r*_, *I*_*c*,2,*r*_,..), making the estimation of the latter based on the former a classical *deconvolution* problem, similar to those known from electronic signal processing, financial time series analysis or medical imaging [13, 14]. For every country, the unknown *I*_*c,w,r*_ were estimated using a deconvolution operation based on maximum likelihood. The total number of new infections among ≥20-year olds during week *w* was estimated as *I*_*c,w*_ = *I*_*c,w,r*_ *∑*_*g*_ *α*_*g*_*N*_*c,g*_*/N*_*c,r*_. Cumulative (i.e., past and current) infection counts were calculated as incremental sums of the weekly infection count estimates. The pandemic’s exponential growth rate over time was subsequently calculated from the estimated weekly infection counts based on a Poisson distribution model and using a sliding-window approach.

Depending on the particular choice of infection risk ratios, this yielded different estimates for the weekly nationwide infection counts, the cumulative infection counts and the exponential growth rates over time. Uncertainty in the true infection risk ratios in any particular country was accounted for by randomly sampling from the full distribution of fitted infection risk ratios multiple times, and calculating confidence intervals of the predictions based on the obtained distribution of estimates. Estimated weekly and cumulative infection fractions (i.e., relative to population size) and exponential growth rates over time are shown for a selection of countries in Fig. 2 and Supplemental Figs. S3 and S4. A comprehensive report of estimates for all 171 countries is provided as Supplemental File 6. Global color-maps of the latest estimates for all countries are shown in Fig. 3.

**Figure 2:**
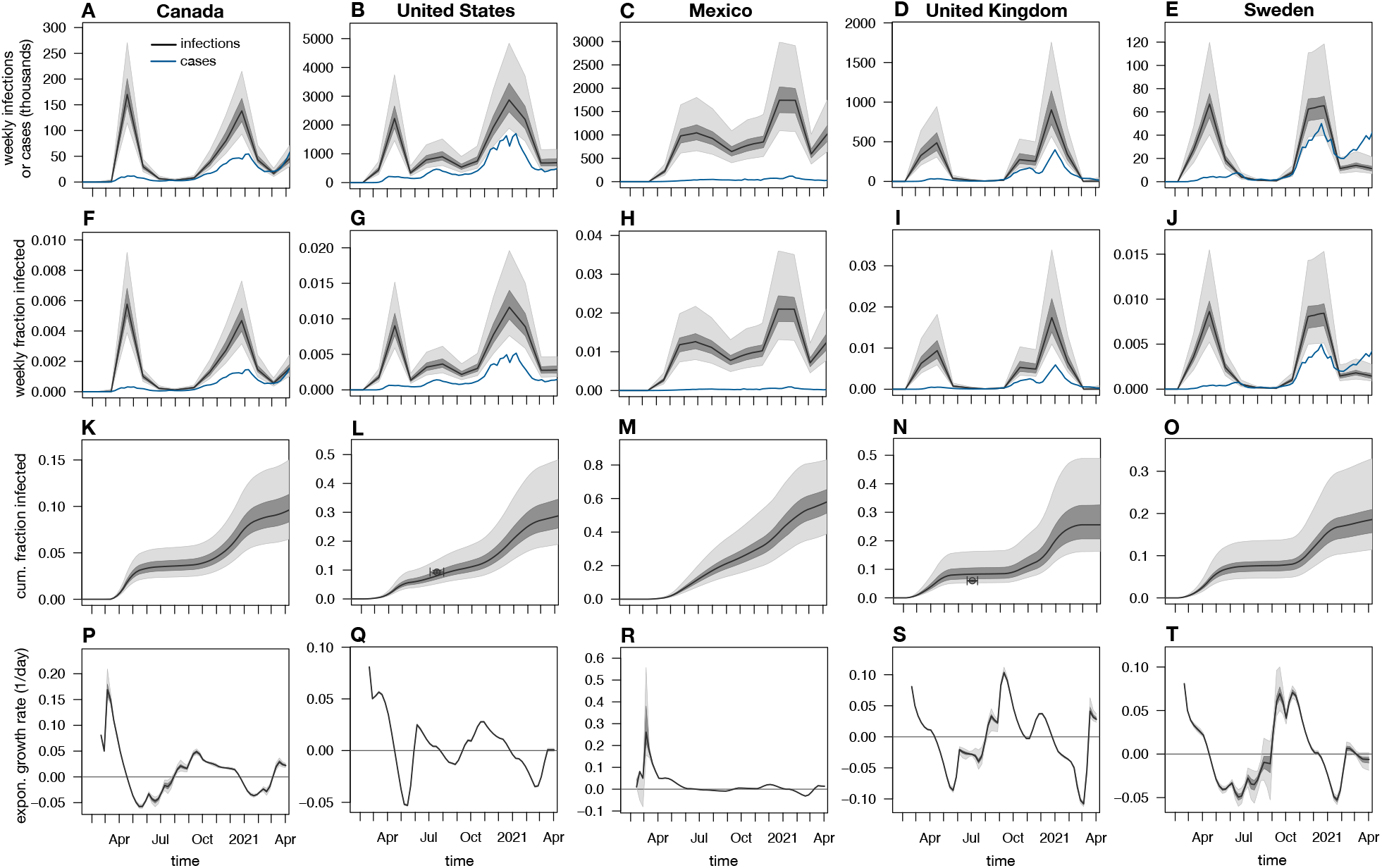
Estimated nationwide infection rates (adults aged ≥20 years). (A–E) Estimated nationwide weekly number of SARS-CoV-2 infections over time, for various countries in North America and Northern Europe, compared to weekly reported cases (blue curves). Black curves show prediction medians, dark and light shades show 50 % and 95 % percentiles of predictions, respectively. Note that reported cases are shown 1 week earlier than actually reported (corresponding roughly to the average incubation time [8]) for easier comparison with infection counts. (F–J) Estimated nationwide weekly fraction of new infections and fraction of reported cases (relative to population size), for the same countries as in A–E. (K–O) Estimated nationwide cumulative fraction of infections (compared to population size), for the same countries as in A–E. Small circles show empirical nationwide prevalence estimates from published seroprevalence surveys for comparison (horizontal error bars denote survey date ranges, vertical error bars denote 95%-confidence intervals as reported by the original publications; references in Supplemental Table S3). (P–T) Estimated exponential growth rate based on weekly infection counts, for the same countries as in A–E. Horizontal axes are shown for reference. Each column shows estimates for a different country. All model estimates refer to adults aged ≥20 years, while reported cases (blue curves) refer to the entire population. The last time point in the plots corresponds to April 9, 2021. Analogous estimates for all 171 investigated countries are provided as Supplemental File 6.

**Figure 3:**
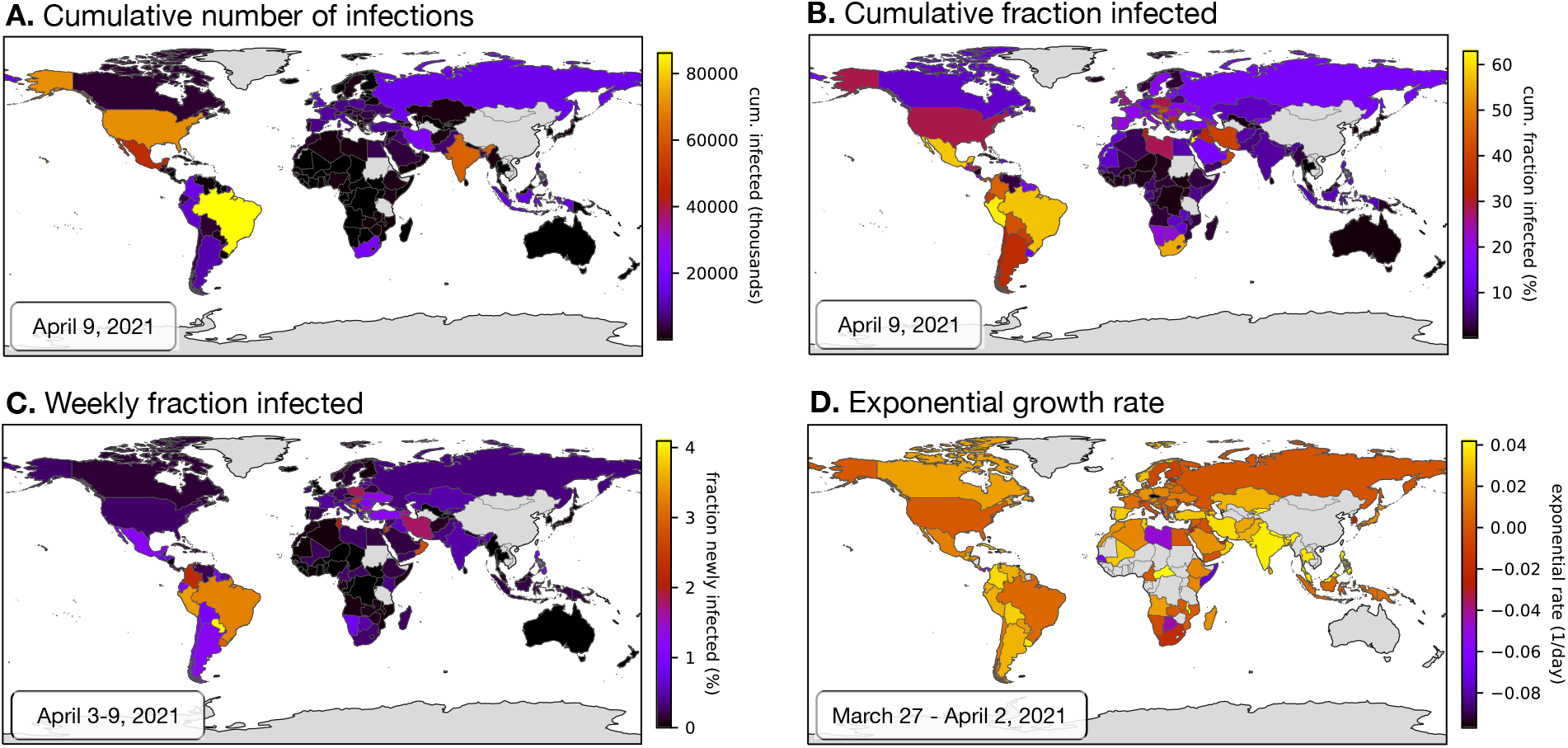
Worldwide overview of latest estimates (adults aged ≥20 years). Global map of the latest estimated nationwide (A) cumulative (past and current) number of SARS-CoV-2 infections, (B) cumulative fraction of infection (infections relative to population size), (C) weekly fraction of new infections (relative to population size) and (D) current exponential growth rate. Dates of the estimations are given in the lower-right corner of each figure. Countries for which an estimation was not performed (e.g., due to insufficient data) are shown in grey. Analogous world maps for older dates are available in Supplemental File 6.

To assess the accuracy of the above approach, I compared the estimated cumulative infection fractions to previously published nationwide antibody-based seroprevalence surveys across 16 countries (Supplemental Table S3). Only surveys attempting to estimate nationwide seroprevalence in the general population (in particular, either using geographically or demographically stratified sampling or adjusting for sample demo-graphics) were included. Agreement between model estimates and seroprevalence estimates was generally good: 16 out of 21 seroprevalence estimates (accounting for the associated 95% confidence interval and the time period of the underlying survey) overlapped with the model’s 95%-confidence intervals, with 3 non-overlaps observed for Brazil, one for India and one for France (Supplemental Fig. S3). Apart from potentially incomplete death count reports causing erroneous model predictions (discussed below), deviations from seroprevalence-based estimates may also be due to the fact that antibody concentrations in infected individuals (especially asymptomatic ones) can drop over time, rendering many of them seronegative [15–17]. Thus, previously infected individuals may not all be recognized as such. Further, sensitivity and specificity estimates for antibody tests performed in the laboratory or claimed by manufactures need not always apply in a community setting [17], thus introducing biases in seroprevalence estimates despite adjustments for sensitivity and specificity.

### Case counts alone can yield wrong impressions

Estimates of the SARS-CoV-2 prevalence in a population can yield insight into the pandemic’s growth dynamics that may not have been possible from reported case counts alone. One reason is that the fraction infections that are detected and reported varies greatly between countries as well as over time. Indeed, according to the present estimates, in most countries case counts initially severely underestimated the actual number of infections and often did not properly reflect the progression of the pandemic, while in many countries more recent case reports capture a much larger fraction of infections and more closely reflect the pandemic’s dynamics (Figs. 2A–E and Supplemental Fig. S4). For example, in the US, France, Sweden, Belgium, Spain, United Kingdom and many other European countries reported cases only reflected a small fraction of infections occurring in Spring 2020, while the majority of subsequent infections have been successfully detected. Nevertheless, in many countries even recent case counts do not correctly reflect the actual dynamics of the pandemic, sometimes even suggesting an opposite trend in its growth. For example, recent reported case counts in Afghanistan, Angola, Brazil, Ecuador, Egypt, Guatemala and Iran severely underestimate the disease’s rapid ongoing growth, with nearly all infections remaining undetected or unreported (Fig. 4). Future investigations, enabled by the infection count estimates presented here, might be able to identify the main factors (e.g., political, financial, organizational) driving the discrepancies between infections and reported cases and suggest concrete steps to eliminate these discrepancies or correct for them.

**Figure 4:**
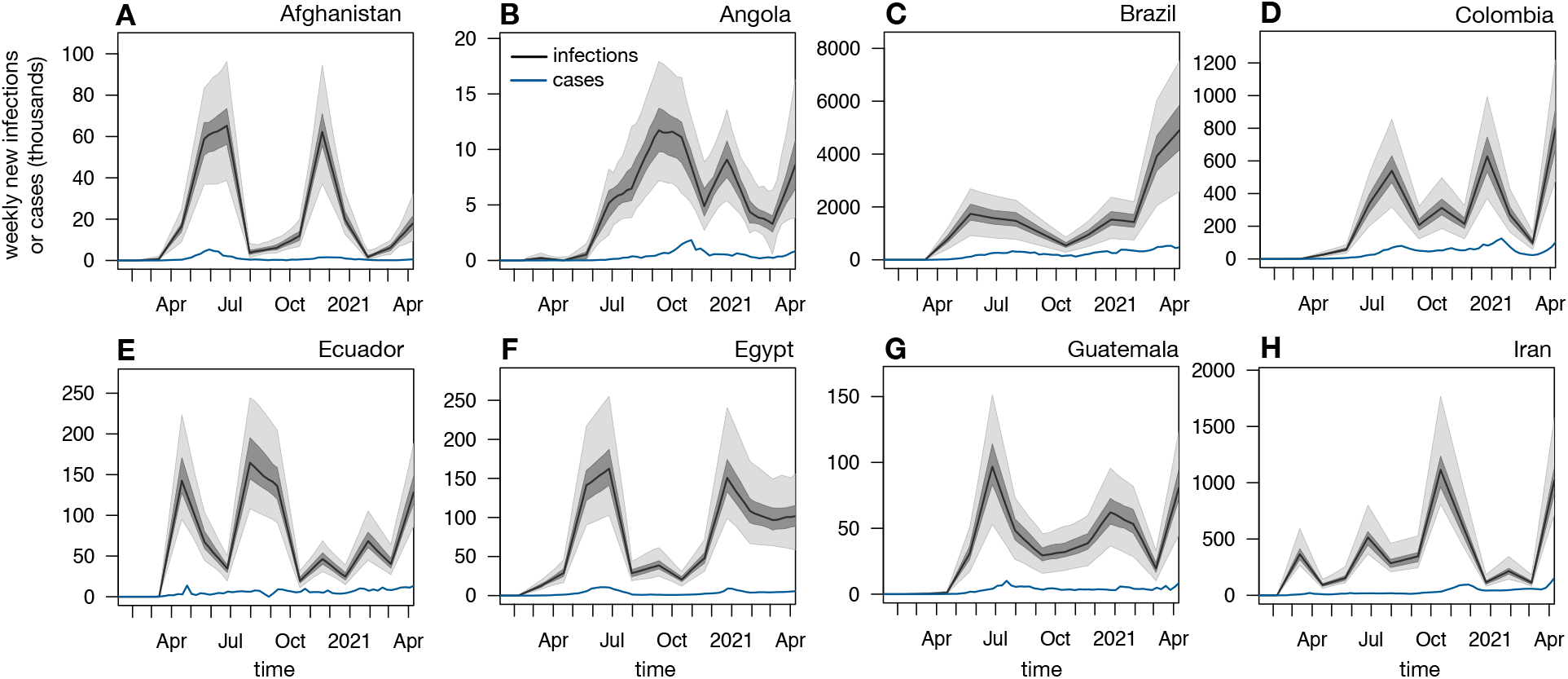
Case counts can suggest drastically different dynamics than infection counts. Nationwide predicted weekly number of new infections (black curves and shades, among adults aged ≥20 years) and weekly reported cases (blue curves, all ages) over time in Afghanistan, Angola, Brazil, Colombia, Ecuador, Egypt, Guatemala and Iran. Black curves show prediction medians, dark and bright shades show 50 % and 95 % confidence intervals, respectively. For easier comparison, case counts are shifted backward by one week (corresponding roughly to the average incubation time [8]).

As infection counts do not depend on testing effort and strategies, they are arguably more suitable than case reports for comparing the pandemic’s progression between countries. For example, my results indicate that as of April 9, 2021 Sweden — often criticized for it’s reluctance to impose strong restrictions on it’s citizens — experienced fewer per-capita infections than many other European countries such as Bulgaria, Czech Republic, Hungary, Poland, Slovakia and Slovenia (Supplemental Fig. S5). Similarly, the highest cumulative per-capita number of cases is reported for the United States, while the estimated fraction of the population actually infected is comparable to many European countries, and about 2 times lower than in Mexico, Peru, Brazil, South Africa or Jordan (Supplemental File 6). These observations highlight the importance of considering actual infection counts (and of course death counts) relative to population size when evaluating policy differences between countries. Future investigations, enabled by the prevalence estimates presented here, may be able to identify concrete political, environmental and socioeconomic factors influencing the pandemic’s growth.

### Caveats

The predictions presented here are subject to some important caveats. First, incomplete, erroneous or age-biased reporting of COVID-19-related deaths will have a direct impact on the estimated infection counts. This caveat is particularly important for countries with less developed medical or reporting infrastructure, as well as for countries were reports may be censored or modified for political reasons. Comparisons of results between countries should thus be done with care. Second, the age-specific infection risk ratios (*α*_*c,g*_) were calibrated based on available age-stratified death statistics from a limited number of countries, and may not apply to all other countries (for example due to strong cultural differences). Uncertainty associated with this extrapolation is partly accounted for by considering infection risk ratios calibrated to multiple alternative countries (see Methods). Third, age-specific IFRs were obtained from studies in only a few countries (mostly western) and often based on a small subset of closely monitored cases (e.g., from the Diamond Princess cruise ship). These IFR estimates may not be accurate for all countries, especially countries with a very different medical infrastructure, different sex ratios in the population or a different prevalence of pre-existing health conditions (e.g., diabetes), all of which can affect the IFR. That said, estimated trends over time within any given country, in particular exponential growth rates (e.g., Figs. 2P–T), are unlikely to be substantially affected by such biases if the biases remain relatively constant over time. For example, the exponential growth rates estimated here remained unchanged when alternative IFRs from the literature [6, 7, 9–12] were considered. To nevertheless examine the robustness of estimated SARS-CoV-2 prevalences against variations in the IFR, I repeated the above analyses by considering for each age group an ensemble of IFRs, i.e., randomly sampling from the set of previously reported IFRs [6, 7, 9–12] rather than considering their mean. Median model predictions remained nearly unchanged, however the uncertainty (i.e., confidence intervals) of the estimates increased (examples in Supplemental Fig. S8).

### Conclusion

I have presented estimates of the nationwide prevalence and growth rate of SARS-CoV-2 infections over time in 171 countries around the world, based on official COVID-19-related death reports, age-specific infection fatality risks, each country’s population age structure and the distribution of time lags between infection, disease onset and death [8]. The complete report for all 171 countries is provided as Supplemental File 6. My estimates are also provided as machine-readable tables (Supplemental Files 1–5) for convenient downstream analyses; periodically-updated estimates are available at: www.loucalab.com/archive/COVID19prevalence. My estimates are largely consistent with data from nationwide general-population seroprevalence surveys. My findings suggest that while in many countries the detection of infections has greatly improved, there are also numerous examples where even recent reported case counts do not properly reflect the pandemic’s dynamics. In particular, comparisons between countries based on infection counts can yield very different conclusions than comparisons merely based on confirmed case counts. My estimates thus enable more precise assessments of the disease’s past and ongoing progression, evaluation and improvement of public interventions and testing strategies, and estimation of worldwide vaccination needs.

## Methods details

### Age-specific infection fatality risks

Age-specific infection fatality risks (IFRs) were calculated based on the following literature: Table 1 in [10], Supplementary Appendix Q in [6], Table S2 in [11], Table 2 in [9], Table S4 in [7], and Eq. (1) in [12]. For each considered age group, the average IFR across all of the aforementioned published IFRs was used, after linearly interpolating where necessary (Supplemental Table S1).

### Calibrating age-specific infection risk ratios

Age-specific population sizes for each country (status 2019) were downloaded from the United Nations website (https://population.un.org/wpp/Download/Standard/CSV) on October 23, 2020 [18]. Time series of nationwide cumulative COVID-19-related death counts grouped by 5-year age intervals were downloaded on April 27, 2021 from COVerAGE-DB (https://osf.io/7tnfh), which is a database that gathers and curates official death count statistics from multiple official sources [19]. The last 7 days covered in the database were ignored to avoid potential biases caused by delays in death reporting. For each country included in COVerAGE-DB, and separately for each age-group, I ensured that cumulative death counts are non-decreasing over time by linearly re-interpolating death counts at problematic time points. The resulting time series were then linearly interpolated onto a regular weekly time grid, i.e., in which adjacent time points are 7 days apart (no extrapolation was performed, i.e., only dates covered by the original time series were included). The weekly number of new deaths in each age group were calculated as the difference of cumulative deaths between consecutive time points on the weekly grid. While some of the input time series are available at a daily resolution, a weekly discretization was chosen here to (a) reduce time series noise and (b) to “average out” the hard-to-model systematic variations in the epidemic’s dynamics between different days of the week (e.g., weekends vs. work days). To ensure a high accuracy in the estimated infection risk ratios, in the following analysis I only considered countries for which COVerAGE-DB covered at least 20 weeks with at least 100 reported deaths each. The following 24 countries were thus considered: Argentina, Austria, Bangladesh, Brazil, Switzerland, Chile, Colombia, Czech Republic, Germany, Spain, France, United Kingdom, Hungary, Indonesia, India, Italy, Mexico, Netherlands, Peru, Philippines, Paraguay, Sweden, Ukraine, United States.

For each considered country *c*, I chose as “reference” age group *r* the age group that had the highest cumulative number of deaths. Designating a reference group is done purely for notational simplicity and consistency, so that age-specific prevalence ratios can all be defined relative to a common reference. For each other age group *g*, I estimated the infection risk ratio *α*_*c,g*_, i.e., the probability of an individual in group *g* being infected relative to the probability of an individual in group *r* being infected, using a probabilistic model according to which the number of deaths in group *g* during week *w* (denoted *D*_*c,w,g*_) was Poisson distributed with expectation:

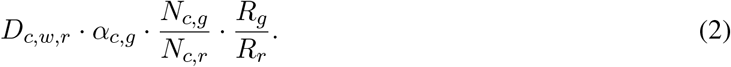

Here, *N*_*c,g*_ is the population size of age group *g* in country *c* and *R*_*g*_ is the IFR for age group *g*. Under this model, the maximum-likelihood estimate for *α*_*c,g*_, i.e. given the weekly death count time series, is given by:

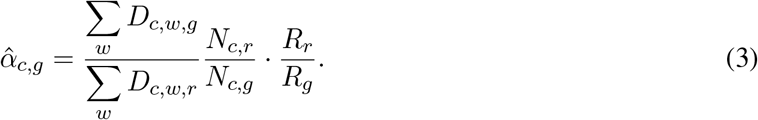

To avoid errors due to sampling noise, only weeks with at least 100 reported deaths were considered in the sums in Eq. (3). I mention that *α*_*c,g*_ might also alternatively be estimated as the slope of the linear regression:

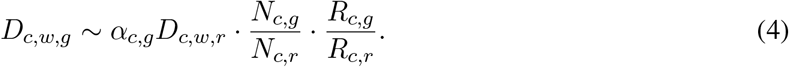

Estimates obtained via linear regression were nearly identical to those obtained using the aforementioned Poissonian model, showing that the estimates were not very sensitive to the precise assumed distribution.

For purposes of evaluating the model’s adequacy (explained below), I also estimated the weekly number of infections in the reference age group, *I*_*c,w,r*_, via maximum-likelihood based on a probabilistic model in which *D*_*c,w,g*_ was Poisson-distributed with expectation:

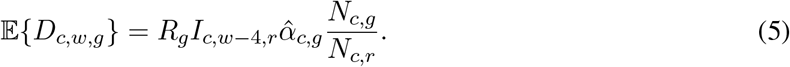

Under this model, the maximum-likelihood estimate for *I*_*c,w−*4,*r*_ is given by:

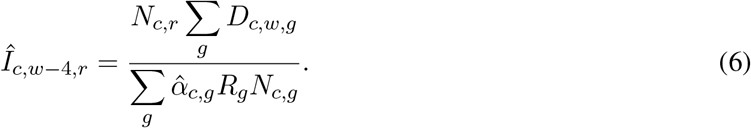

To evaluate the adequacy of the above stochastic model in explaining the original death count data, I simulated multiple hypothetical weekly death counts for each age group and compared the distribution of simulated death counts to the true death counts. Specifically, for each country *c*, week *w* and age group *g*, I drew 100 random death counts 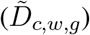 from a Poisson distribution with expectation:

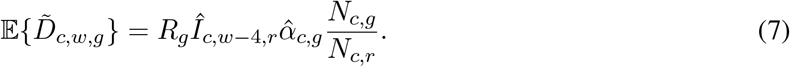

Median simulated death counts and 50 % and 95% confidence intervals, along with the original death counts, are shown for a representative selection of countries and age groups in Supplemental Fig. S2. As can be seen in that figure, the model’s simulated time series are largely consistent with the original data.

### Estimating infection counts from total death counts

Time series of total (non-age-stratified) nationwide cumulative reported death and case counts were downloaded from the website of the World Health Organization (https://covid19.who.int/table) on May 1, 2021. The last 7 days covered in the database were ignored to avoid potential biases caused by delays in case and death reporting. Cumulative death and case counts were made non-decreasing and interpolated onto a weekly time grid as described above. Only countries that reported at least one death per week for at least 10 weeks were included in the analysis below. For each country *c*, week *w* and any particular choice of age-specific infection risk ratios *α*_1_, *α*_2_,.., the number of infections was estimated as follows. Let *N* be the number of consecutive weeks for which total deaths are reported. Let *r* denote some fixed reference age group with respect to which infection risk ratios are defined, i.e., such that *α*_*r*_ = 1 (here, ages 70–74 were used as reference). Let *δ*_*k*_ denote the probability that a fatal infection will lead to death after *k* weeks, where *k* = *L*_min_,.., *L*_max_ and where *L*_min_ is the minimum and *L*_max_ the maximum considered time lag. Let *L* := *L*_max_ − *L*_min_ + 1. Let *I*_*c,w,r*_ be the (a priori unknown) number of new infections occurring during week *w* in the reference age group. The number of COVID-19-induced deaths during week *w* in age group *g*, denoted *D*_*c,w,g*_, was assumed to be Poisson-distributed with expectation equal to:

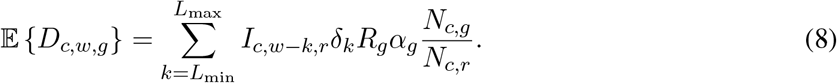

The total number of deaths in week *w, D*_*c,w*_, is thus Poisson-distributed with expectation:

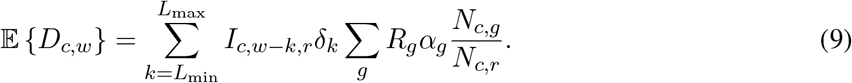

As explained in the main text, only age groups ≥20 years were included because infection risk ratios could not be reliably estimated for younger ages and because the contribution of younger ages to total death counts can be considered numerically negligible. Here, the *δ*_*k*_ were calculated using 1,000,000 Monte Carlo simulations based on the log-normal distribution models fitted by Linton *et al*. [8, Table 2 therein] for the time lags between infection and disease onset and the time lags between disease onset and death, and assuming that the two time lags are independently distributed (see Supplemental Table S2). The minimum and maximum considered lags were *L*_min_ = 2 and *L*_max_ = 6 weeks, since this range covers the bulk (~90%) of cases, and since further increasing increasing *L*_max_ or decreasing *L*_min_ increases the width of the convolution kernel, thus increasing the risk of introducing spurious fluctuations in the estimated *I*_*c,w,r*_.

Given the above model, our goal is to estimate the unknown weekly infection counts in the reference group, *I*_*c,w,r*_ from the recorded weekly death counts *D*_*c,w*_. Note that this is a classical deconvolution problem, since each *D*_*c,w*_ results from the additive effects of infections from multiple preceding weeks. [13, 14]. Eq. (9) can be written abstractly in matrix form:

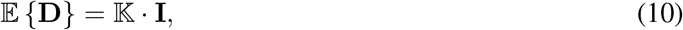

where 𝕂 is a *convolution matrix* of size *N* × (*N* + *L* − 1):

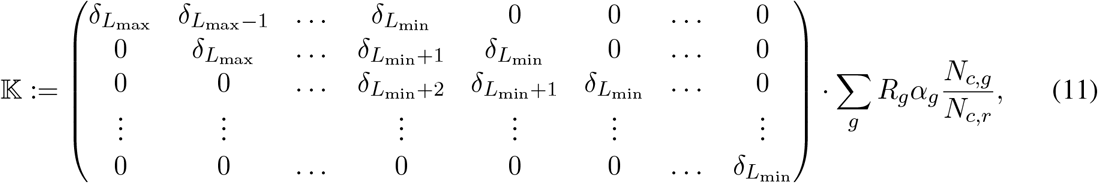

and **D** is a column vector of size *N* listing the reported weekly death counts *D*_*c*,1_,.., *D*_*c,N*_ and **I** is a column vector of size *N* + *L* −1 listing the unknown weekly infection counts 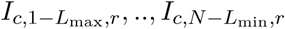. Note that I omitted the country index *c* from the **I, D**, 𝕂 for notational simplicity, but keep in mind that **I, D**, 𝕂 refer to a specific country. It is straightforward to show that, under the above model, the log-likelihood of the observed weekly death counts (**D**) is given by:

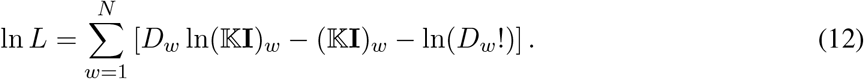

In principle, one could estimate the unknown vector **I** via maximum-likelihood. Indeed, the above log-likelihood is maximized when the following condition is met:

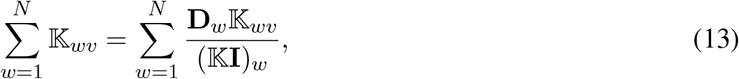

for all *v* ∈{1,.., *N* + *L* −1}. A sufficient condition for Eq. (13) is that 𝕂**I** = **D**, in other words any vector **Î** satisfying 𝕂**Î** = **D** is a maximum-likelihood estimate. Such an estimate can be obtained using the Moore-Penrose pseudoinverse of 𝕂, denoted 𝕂^+^ [20, 21]: Since 𝕂 has linearly independent rows, its pseudoinverse is 𝕂^+^ = 𝕂^T^(𝕂 𝕂^T^)^−1^, and hence setting **Î** := 𝕂^+^**D** would satisfy 𝕂**Î** = **D**. However, due to known issues with inverting convolution matrixes such a naive estimation tends to introduce spurious fluctuations in the estimated **I**. One approach is to reduce the temporal resolution of the estimated **I**, which effectively reduces the number of estimated free parameters [22]. Hence, instead of estimating *I*_*c,w,r*_ separately for each week, I considered a coarser time grid that has 4 fewer time points than the original weekly time grid, i.e., such that the infection count *I*_*c,w,r*_ is freely estimated only every 4-th week, while assuming linear variation between these time points. For example, for an original weekly time series spanning 100 weeks, I first estimated the *I*_*c,w,r*_ at about 100*/*4 discrete time points, each 4 weeks apart, and then used linear interpolation to obtain the remaining *I*_*c,w,r*_. Denoting by **J** the vector listing the infection counts on this coarser time grid 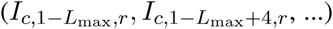, and by 𝔾 the matrix mapping **J** to **I** via linear interpolation (i.e., **I** = 𝔾 **J**), we thus obtain the following log-likelihood in terms of **J** :

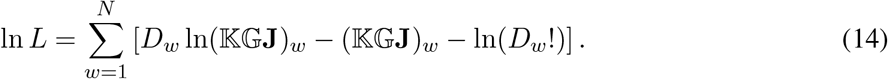

The corresponding-maximum likelihood estimate **Ĵ** can no longer be obtained simply by solving the equation 𝕂 𝔾 **Ĵ** = **D**, because this linear problem is over-determined, i.e., it is unlikely that a **Ĵ** can be found such that 𝕂 𝔾 **Ĵ** = **D** is exactly satisfied. However an optimally approximate solution (in the least-squares sense), 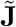, can be obtained by setting 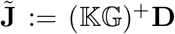. In order to determine the exact maximum-likelihood estimate **Ĵ**, i.e., the **J** maximizing ln *L* in Eq. (14), I used numerical optimization, implemented in the R function nloptr::nloptr, while using 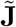 as a starting point. Subsequently setting **Î** := 𝔾 **Ĵ** yielded an estimate for the weekly infections counts *I*_*c,w,r*_. The corresponding total number of weekly infections, *Î*_*c,w*_, can be calculated from the estimates *Î*_*c,w,r*_ as follows:

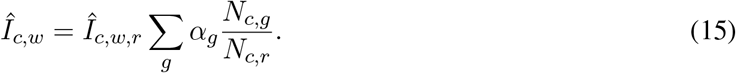

The corresponding cumulative number of total infections up until any given week can be obtained by summing the weekly infection counts.

Exponential growth rates over time were estimated from the weekly infection counts using a sliding-window approach, as follows. In every sliding window (spanning 5 consecutive weeks), an exponential function of the form *I*(*t*) = *Ae*^*tλ*^ was fitted, where *t* denotes time in days and *A* and *λ* are unknown parameters (in particular, *λ* is the exponential growth rate in that window). The parameters *A* and *λ* were fitted via maximum likelihood, assuming that the total number of weekly infections, *I*_*c,w*_, was Poisson distributed with expectation 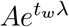. Under this model, the log-likelihood of the data (more precisely, of the previously estimated weekly infection counts) is:

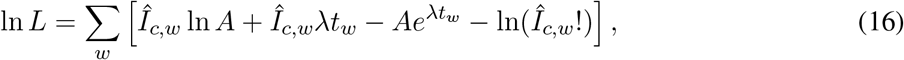

where *w* iterates over all weeks in the specific sliding window. The maximum-likelihood estimates of *A* and *λ* are obtained by solving *∂* ln *L/∂λ* = 0 and *∂* ln *L/∂A* = 0, which quickly leads to the condition:

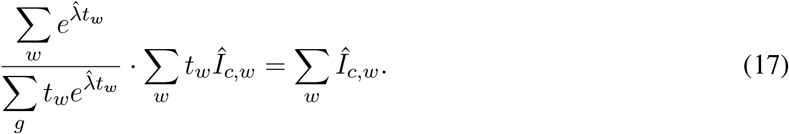

Equation (17) was solved numerically to obtain the maximum-likelihood estimate 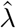.

To assess estimation uncertainties stemming from sampling stochasticity and uncertainties in the infection risk ratios, I repeated the above estimations 100 times using alternative infection risk ratios (for each age group drawn randomly from the set of infection risk ratios previously fitted to various countries) and replacing the reported weekly death counts *D*_*c,w*_ with values drawn from a Poisson distribution with mean *D*_*c,w*_. Hence, rather than point-estimates, all predictions are reported in the form of medians and confidence intervals. Only infection risk ratios for which the corresponding linear curve (Eq. 4) achieved a coefficient of determination (*R*^2^) greater than 0.5 were used (shown in Fig. 1), to avoid less accurately estimated infection risk ratios (typically obtained from countries with low death rates). Tables of all estimates for all considered countries up until April 9, 2021 are provided in Supplemental Files 1–5; a visual report is provided as Supplemental File 6.

## Data Availability

All data used in this manuscript were obtained from publicly accessible sources.

https://covid19.who.int/table

https://population.un.org/wpp/Download/Standard/CSV

https://osf.io/7tnfh

## Data availability

All data used in this manuscript are publicly available at the locations described in the Methods section.

## Competing interests

The author declares no conflict of interest.

## Acknowledgements

The author was supported by a US National Science Foundation RAPID grant.

## Supplementary Information

**Table S1:** Age-specific infection fatality risks used in this study, obtained by averaging IFRs reported in multiple previous studies (see Methods for details and sources).

| age group (years) | IFR (%) |
| --- | --- |
| 0–4 | 0.0015 |
| 5–9 | 0.00257 |
| 10–14 | 0.0102 |
| 15–19 | 0.0102 |
| 20–24 | 0.0115 |
| 25–29 | 0.0143 |
| 30–34 | 0.0221 |
| 35–39 | 0.0384 |
| 40–44 | 0.0641 |
| 45–49 | 0.118 |
| 50–54 | 0.209 |
| 55–59 | 0.403 |
| 60–64 | 0.739 |
| 65–69 | 1.41 |
| 70–74 | 2.56 |
| 75–79 | 5.55 |
| 80–84 | 9.47 |
| 85–89 | 10.77 |
| $\geq 90$ | 11.99 |

**Table S2:** Discretized probability distribution of the time lag between infection and COVID-19-induced death, as used in this paper, calculated based on the log-normal models by [8]. Specifically, *δ*_*k*_ was calculated as the probability of death occurring between 7 *δ*_*k*_ − 3.5 and 7 *δ*_*k*_ + 3.5 days from infection, conditioned on the infection being fatal, with all *δ*_*k*_ subsequently normalized so that their sum is 1. Note that deaths are also possible at shorter or longer time delays, albeit at much lower probabilities.

| $k$ | $\delta_k$ |
| --- | --- |
| 2 | 0.250 |
| 3 | 0.323 |
| 4 | 0.229 |
| 5 | 0.130 |
| 6 | 0.068 |

**Table S3:** Previously published nationwide seroprevalence estimates, considered in this study for comparison (Supplemental Figs. S3, S6, S8). Also listed are the start and end dates of the underlying surveys.

| country | start date | end date | seroprevalence (95% CI) | reference |
| --- | --- | --- | --- | --- |
| Brazil | 2020-05-01 | 2020-05-15 | 0.014 (0.013–0.016) | [23] |
| Brazil | 2020-05-14 | 2020-05-21 | 0.019 (0.017–0.022) | [24] |
| Brazil | 2020-06-04 | 2020-06-07 | 0.031 (0.028–0.034) | [24] |
| Denmark | 2020-08-01 | 2020-08-31 | 0.02 (0.017–0.024) | [25] |
| Denmark | 2020-10-01 | 2020-10-31 | 0.024 (0.019–0.028) | [25] |
| Denmark | 2020-12-01 | 2020-12-31 | 0.043 (0.034–0.051) | [25] |
| France | 2020-03-09 | 2020-03-15 | 0.0041 (0.0005–0.0088) | [26] |
| France | 2020-04-06 | 2020-04-12 | 0.0414 (0.031–0.0499) | [26] |
| France | 2020-05-11 | 2020-05-17 | 0.0493 (0.0402–0.0589) | [26] |
| Greece | 2020-03-01 | 2020-03-31 | 0.0002 (0.00–0.00025) | [27] |
| Greece | 2020-04-01 | 2020-04-30 | 0.0025 (0.0002–0.005) | [27] |
| Hungary | 2020-05-01 | 2020-05-16 | 0.0068 (0.005–0.0086) | [28] |
| India | 2020-05-11 | 2020-06-04 | 0.0073 (0.0034–0.0113) | [29] |
| India | 2020-08-18 | 2020-09-20 | 0.066 (0.058–0.074) | [30] |
| Israel | 2020-06-28 | 2020-09-14 | 0.038 (0.037–0.040) | [31] |
| Luxemburg | 2020-04-16 | 2020-05-05 | 0.020586 (0.0134–0.0277) | [32] |
| Saudi Arabia | 2020-06-01 | 2020-11-30 | 0.109 (0.103–0.116) | [33] |
| Slovenia | 2020-04-20 | 2020-05-01 | 0.0093 (0.00–0.0265) | [34] |
| Spain | 2020-04-27 | 2020-05-11 | 0.046 (0.043–0.050) | [35] |
| United Kingdom | 2020-06-20 | 2020-07-13 | 0.060 (0.058–0.061) | [36] |
| United States | 2020-07-01 | 2020-07-31 | 0.093 (0.088–0.099) | [37] |

**Figure S1:**
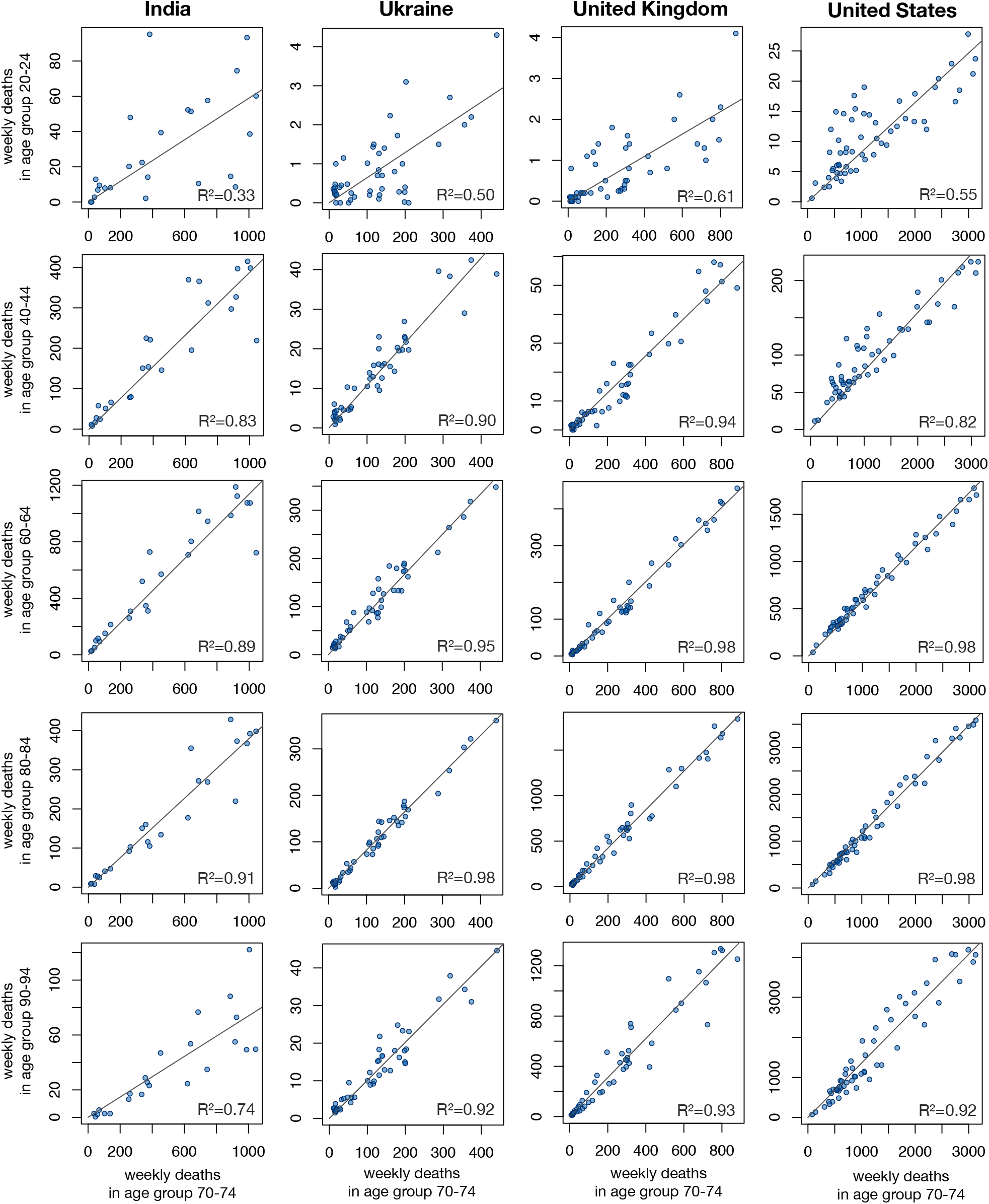
Comparison of weekly death counts between age groups. Weekly COVID-19-related death counts per age group (vertical axes) compared to death counts in the same week in age group 70–74 (horizontal axes), in various countries for which age-stratified death counts were available. Each column corresponds to a different country, each row to a different age group, and each point to a specific week. Least-squares regression lines (with zero intercept) are shown for reference; the corresponding fractions of explained variance (*R*^2^) are shown in the figures.

**Figure S2:**
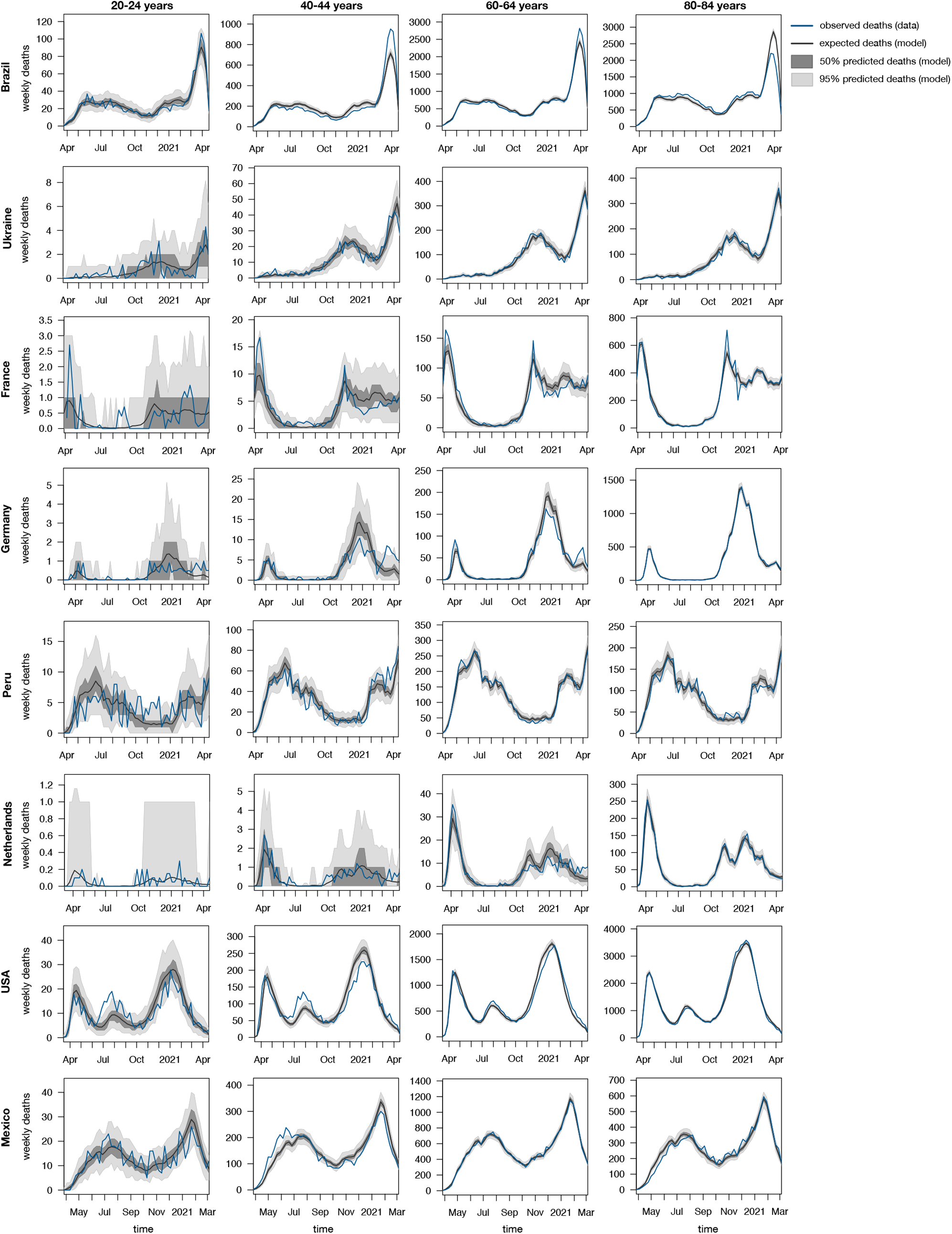
Reported weekly death counts for various countries and selected age groups (blue curves, source: COVerAGE database), compared to death counts predicted by the stochastic model fit to age-stratified death counts (black curves are expectations, dark shades denote 50% and light shades denote 95% confidence intervals, see Eqs. 5–7 in the Methods). Each sub-figure shows a distinct age group for a distinct country.

**Figure S3:**
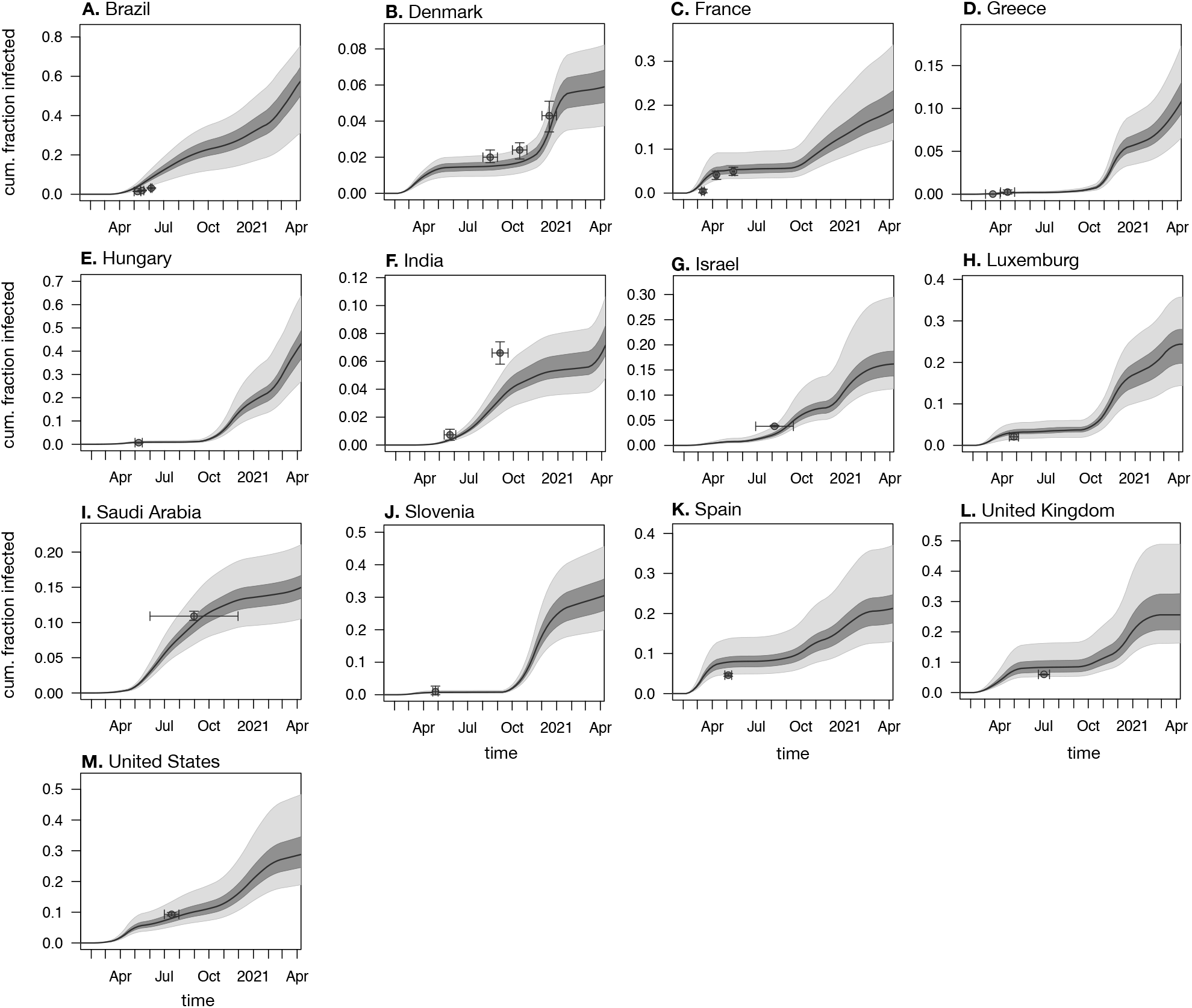
Comparison of predicted infection fractions to seroprevalence data. Predicted cumulative infection fractions relative to population size (aka. prevalence) in various countries (among adults aged ≥20 years). Black curves show prediction medians, dark and light shades show 50 % and 95 % confidence intervals, respectively. Small circles show empirical nationwide prevalence estimates from published seroprevalence surveys for comparison (horizontal error bars denote survey date ranges, vertical error bars denote 95%-confidence intervals as reported by the original publications). Seroprevalence data sources are listed in Supplemental Table S3.

**Figure S4:**
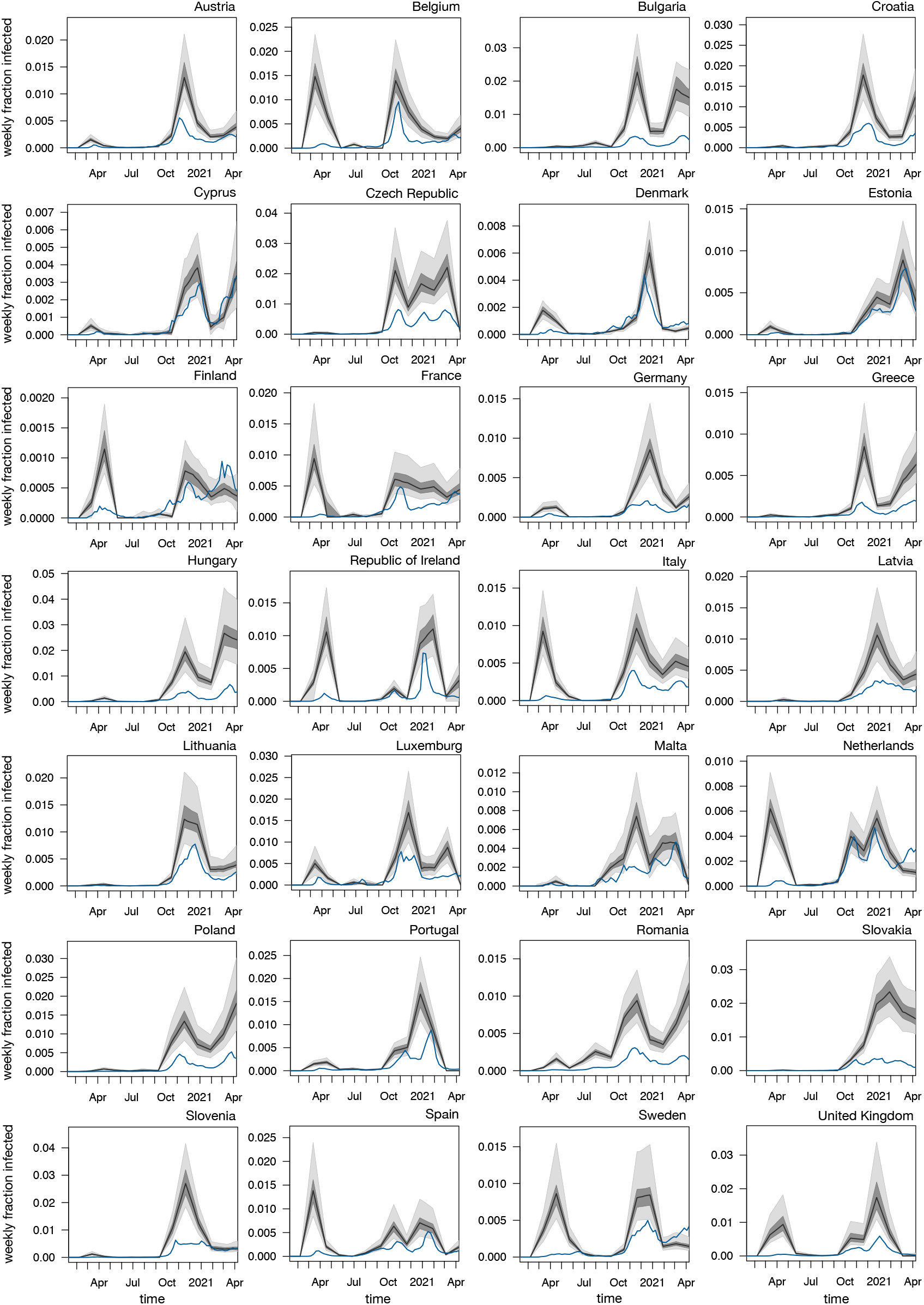
Weekly infection fractions in Europe. Estimated weekly nationwide fraction of new infections (relative to population size) over time, in member countries of the European Union as well as the United Kingdom, among adults aged ≥20 years. Black curves show prediction medians, dark and bright shades show 50 % and 95 % confidence intervals, respectively. Blue curves show weekly reported case fractions (all ages). Note that cases are shown 1 week earlier than actually reported (corresponding roughly to the average incubation time [8]) for easier comparison with infection counts. For cumulative infection fractions see Supplemental Fig. S5.

**Figure S5:**
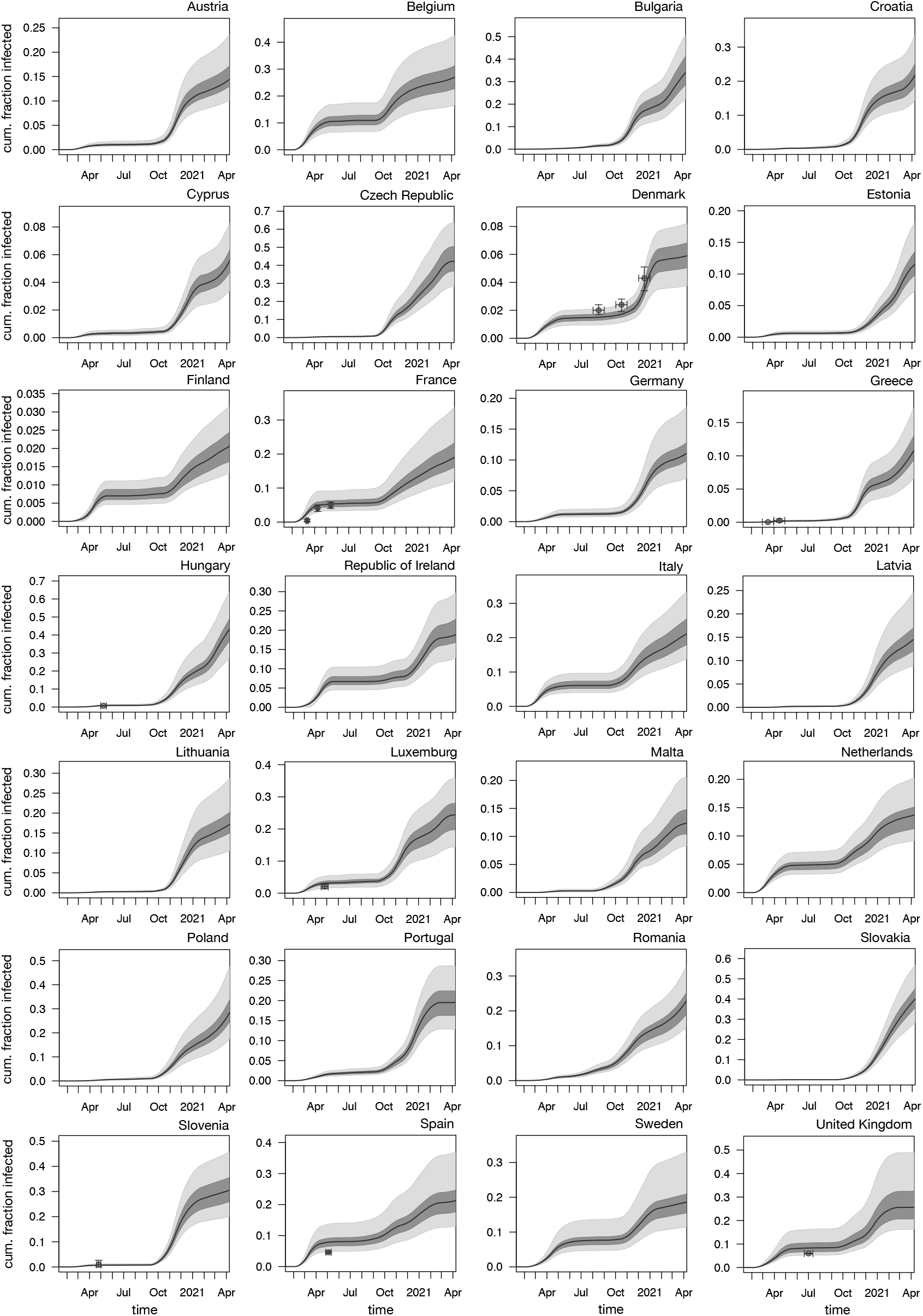
Cumulative infection fractions in Europe. Estimated cumulative nationwide fraction of infections (relative to population size) over time, in member countries of the European Union as well as the United Kingdom, among adults aged ≥20 years. Black curves show prediction medians, dark and bright shades show 50 % and 95 % confidence intervals, respectively. Dots in some figures show estimates from seroprevalence surveys (listed in Table S3). For weekly infection fractions see Supplemental Fig. S4.

**Figure S6:**
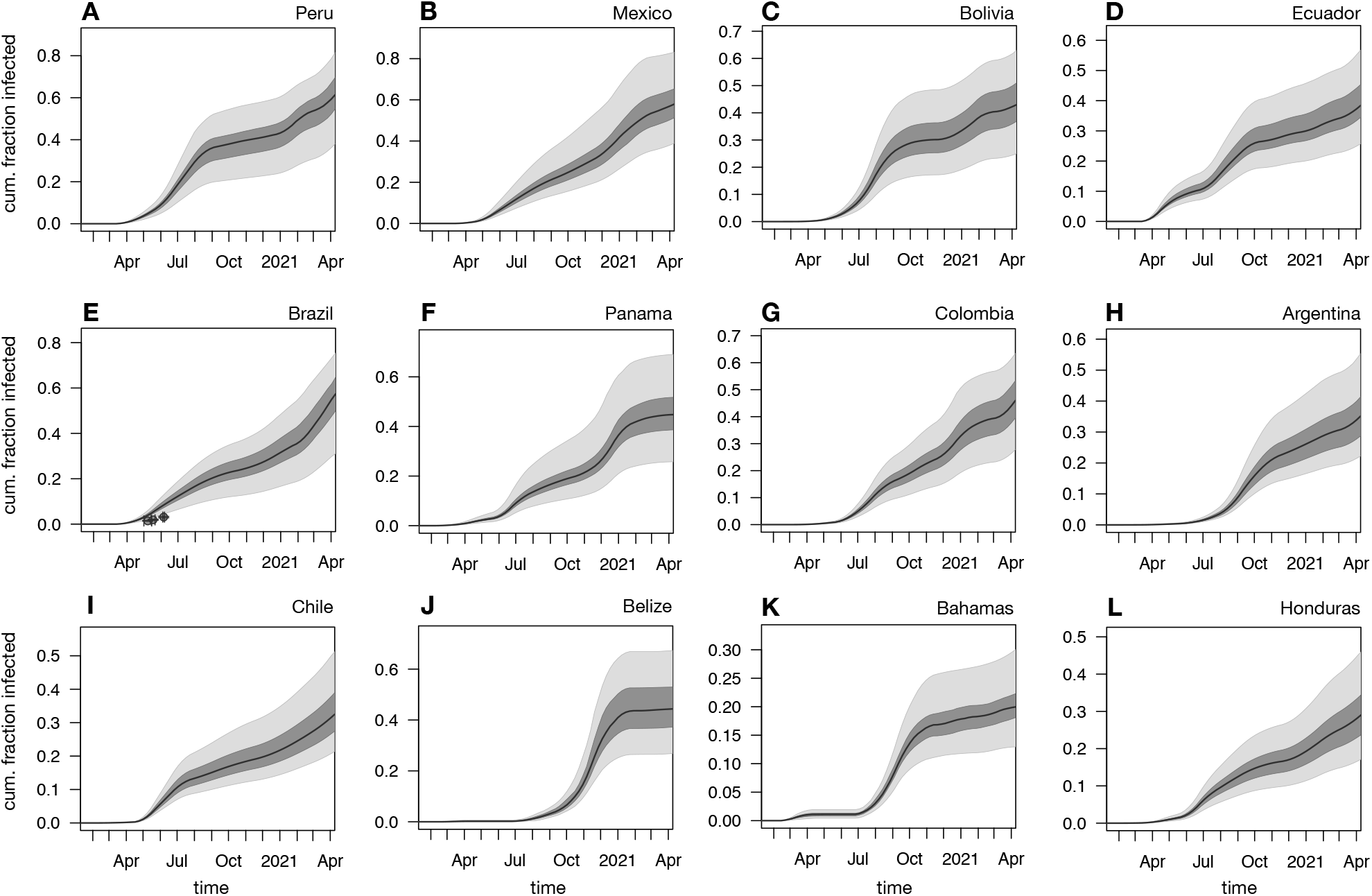
Cumulative infection fractions in the Americas. Estimated nationwide cumulative fraction of infections (relative to population size) over time, among adults aged ≥20 years, in countries of the Americas with particularly high estimates of infections. Black curves show prediction medians, dark and bright shades show 50 % and 95 % confidence intervals, respectively. Dots in (E) show estimates from seroprevalence surveys (listed in Table S3). For weekly infection fractions see Supplemental Fig. S7.

**Figure S7:**
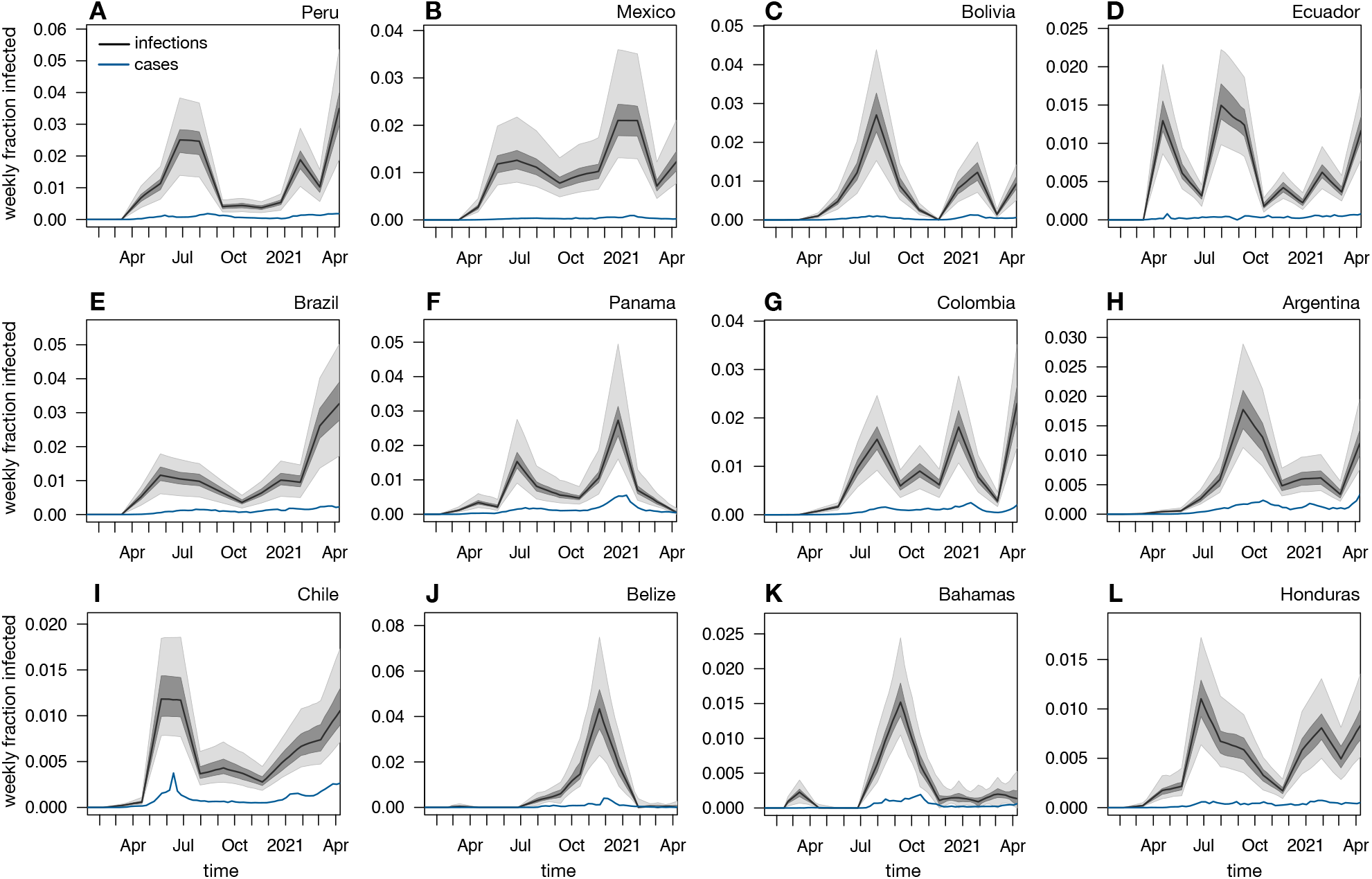
Weekly infection fractions in the Americas. Estimated nationwide weekly fraction of infections (relative to population size) over time, among adults aged ≥20 years, in countries of the Americas with particularly high estimates of infections. Black curves show prediction medians, dark and bright shades show 50 % and 95 % confidence intervals, respectively. Blue curves show weekly reported case fractions (all ages), shifted back by 1 week (corresponding roughly to the average incubation time [8]) for easier comparison with infection counts. For cumulative infection fractions see Supplemental Fig. S6.

**Figure S8:**
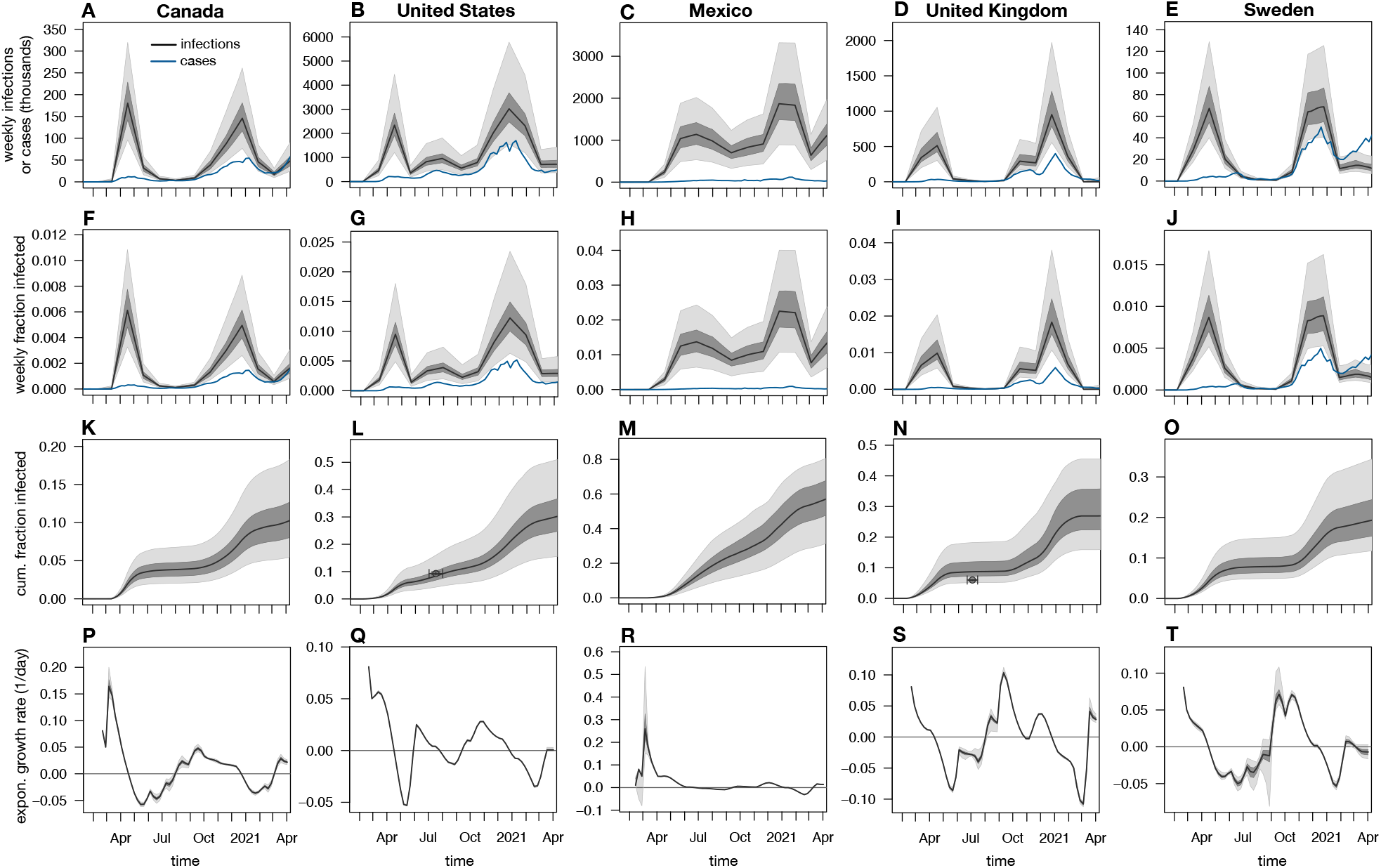
Estimated nationwide infection counts (using the IFR ensemble). Estimated nationwide weekly infection counts, infection fractions (relative to population), cumulative fractions infected and exponential growth rates, among adults aged ≥20 years, for the same countries as in Fig. 2 in the main article. Black curves show prediction medians, dark and light shades show 50 % and 95 % percentiles of predictions, respectively. Blue curves show weekly numbers of reported cases (1st row) or weekly fractions of reported cases (relative to the population size, 2nd row). Note that reported cases are shown 1 week earlier than actually reported (corresponding roughly to the average incubation time [8]) for easier comparison with infection counts. Estimates take into account the variability of IFRs reported in the literature. Dots in L and N show estimates from seroprevalence surveys (listed in Table S3).

## Notes

### Competing Interest Statement

The authors have declared no competing interest.

### Funding Statement

This work was supported by a US National Science Foundation RAPID grant.

### Summary of Updates

Improved methods; updated estimates to cover more recent dates; updated discussion.

## References

[1] Pearce, N., Vandenbroucke, J.P., VanderWeele, T.J. & Greenland, S. Accurate statistics on COVID-19 are essential for policy guidance and decisions. American Journal of Public Health 110, 949–951 (2020).

[2] Lachmann, A., Jagodnik, K.M., Giorgi, F.M. & Ray, F. Correcting under-reported COVID-19 case numbers: estimating the true scale of the pandemic. medRxiv 2020.03.14.20036178 (2020).

[3] Lu, F.S. et al.. Estimating the cumulative incidence of COVID-19 in the United States using four complementary approaches. medRxiv (2020).

[4] Flaxman, S. et al. Estimating the effects of non-pharmaceutical interventions on COVID-19 in Europe. Nature 584, 257–261 (2020).

[5] Dowd, J.B. et al. Demographic science aids in understanding the spread and fatality rates of COVID-19. Proceedings of the National Academy of Sciences 117, 9696–9698 (2020).

[6] Levin, A.T. et al. Assessing the age specificity of infection fatality rates for COVID-19: Meta-analysis & public policy implications. Working Paper 27597, National Bureau of Economic Research (2020).

[7] O’Driscoll, M. et al. Age-specific mortality and immunity patterns of SARS-CoV-2. Nature 590, 140–145 (2021).

[8] Linton, N.M. et al. Incubation period and other epidemiological characteristics of 2019 novel coronavirus infections with right truncation: A statistical analysis of publicly available case data. Journal of Clinical Medicine 9 (2020).

[9] Rinaldi, G. & Paradisi, M. An empirical estimate of the infection fatality rate of COVID-19 from the first Italian outbreak. medRxiv (2020).

[10] Pastor-Barriuso, R. et al. Infection fatality risk for sars-cov-2 in community dwelling population of spain: nationwide seroepidemiological study. BMJ 371, m4509 (2020).

[11] Salje, H. et al. Estimating the burden of SARS-CoV-2 in France. Science 369, 208–211 (2020).

[12] Linden, M. et al. The foreshadow of a second wave: An analysis of current COVID-19 fatalities in Germany. arXiv (2020).

[13] Wiener, N. Extrapolation, Interpolation, and Smoothing of Stationary Time Series (MIT Press, Cambridge, Massachusetts, 1964).

[14] Mendel, J.M. Maximum-Likelihood Deconvolution (Springer, New York, 1990).

[15] Long, Q.X. et al. Clinical and immunological assessment of asymptomatic SARS-CoV-2 infections. Nature Medicine 26, 1200–1204 (2020).

[16] Bolotin, S. et al. SARS-CoV-2 seroprevalence survey estimates are affected by anti-nucleocapsid antibody decline. medRxiv (2020).

[17] La Marca, A. et al. Testing for SARS-CoV-2 (COVID-19): a systematic review and clinical guide to molecular and serological in-vitro diagnostic assays. Reproductive BioMedicine Online 41, 483–499 (2020).

[18] DESA, U. World population prospects 2019, online edition. rev. Tech. Rep., United Nations, Department of Economic and Social Affairs, Population Division (2019).

[19] Riffe, T. et al. Coverage-db: A database of age-structured covid-19 cases and deaths. medRxiv (2020).

[20] Moore, E.H. On the reciprocal of the general algebraic matrix. Bulletin of the American Mathematical Society 26, 394–395 (1920).

[21] Penrose, R. A generalized inverse for matrices. Proceedings of the Cambridge Philosophical Society 51, 406–413 (1955).

[22] Louca, S., Astor, Y.M., Doebeli, M., Taylor, G.T. & Scranton, M.I. Microbial metabolite fluxes in a model marine anoxic ecosystem. Geobiology 17, 628–642 (2019).

[23] Hallal, P. et al. Remarkable variability in SARS-CoV-2 antibodies across Brazilian regions: nationwide serological household survey in 27 states. medRxiv (2020).

[24] Hallal, P.C. et al. SARS-CoV-2 antibody prevalence in Brazil: results from two successive nationwide serological household surveys. The Lancet Global Health 8, e1390–e1398 (2020).

[25] Espenhain, L. et al. Prevalence of SARS-CoV-2 antibodies in Denmark 2020: results from nationwide, population-based sero-epidemiological surveys. medRxiv (2021).

[26] Vu, S.L. et al. Prevalence of SARS-CoV-2 antibodies in France: results from nationwide serological surveillance. medRxiv (2020).

[27] Bogogiannidou, Z. et al. Repeated leftover serosurvey of SARS-CoV-2 IgG antibodies, Greece, March and April 2020. Eurosurveillance 25 (2020).

[28] Merkely, B. et al. Novel coronavirus epidemic in the Hungarian population, a cross-sectional nationwide survey to support the exit policy in Hungary. GeroScience 42, 1063–1074 (2020).

[29] Murhekar, M. et al. Prevalence of SARS-CoV-2 infection in India: Findings from the national serosurvey, May-June 2020. Indian Journal of Medical Research 152, 48–60 (2020).

[30] Murhekar, M.V. et al. Sars-cov-2 antibody seroprevalence in india, august–september, 2020: findings from the second nationwide household serosurvey. The Lancet Global Health 9, e257–e266 (2021).

[31] Reicher, S. et al. Nationwide seroprevalence of antibodies against SARS-CoV-2 in Israel. European Journal of Epidemiology (2021).

[32] Snoeck, C.J. et al. Prevalence of SARS-CoV-2 infection in the Luxembourgish population: the CONVINCE study. medRxiv (2020).

[33] Alharbi, N.K. et al. Nationwide seroprevalence of SARS-CoV-2 in Saudi Arabia. Journal of Infection and Public Health (2021).

[34] Poljak, M. et al. Seroprevalence of severe acute respiratory syndrome coronavirus 2 in slovenia: results of two rounds of a nationwide population study on a probability-based sample, challenges and lessons learned. Clinical Microbiology and Infection (2021).

[35] Pollán, M. et al. Prevalence of SARS-CoV-2 in Spain (ENE-COVID): a nationwide, population-based seroepidemiological study. The Lancet 396, 535–544 (2020).

[36] Ward, H. et al. Antibody prevalence for SARS-CoV-2 in England following first peak of the pandemic: REACT2 study in 100,000 adults. medRxiv (2020).

[37] Anand, S. et al. Prevalence of SARS-CoV-2 antibodies in a large nationwide sample of patients on dialysis in the USA: a cross-sectional study. The Lancet 396, 1335–1344 (2020).

